# Estimating the direct effects of the genetic liabilities to bipolar disorder, schizophrenia, and behavioral traits on suicide attempt using a multivariable Mendelian randomization approach

**DOI:** 10.1101/2023.08.14.23294083

**Authors:** Brenda Cabrera-Mendoza, Necla Aydin, Gabriel R. Fries, Anna R. Docherty, Consuelo Walss-Bass, Renato Polimanti

## Abstract

Bipolar disorder (BD) and schizophrenia (SZ) are associated with higher odds of suicide attempt (SA). In this study, we aimed to explore the effect of BD and SZ genetic liabilities on SA, also considering the contribution of behavioral traits, socioeconomic factors, and substance use disorders. Leveraging large-scale genome-wide association data from the Psychiatric Genomics Consortium (PGC) and the UK Biobank (UKB), we conducted a two-sample Mendelian randomization (MR) analysis to evaluate the putative causal effect of BD (41,917 cases, 371,549 controls) and SZ (53,386 cases, 77,258 controls) on SA (26,590 cases, 492,022 controls). Then, we assessed the putative causal effect of BD and SZ on behavioral traits, socioeconomic factors, and substance use disorders. Considering the associations identified, we evaluated the direct causal effect of behavioral traits, socioeconomic factors, and substance use disorders on SA using a multivariable MR approach. The genetic liabilities to BD and SZ were associated with higher odds of SA (BD odds ratio (OR)=1.24, p=3.88×10^−12^; SZ OR=1.09, p=2.44×10^−20^). However, while the effect of mental distress (OR=1.17, p=1.02×10^−4^) and risk-taking (OR=1.52, p=0.028) on SA was independent of SZ genetic liability, the BD-SA relationship appeared to account for the effect of these risk factors. Similarly, the association with loneliness on SA was null after accounting for the effect of SZ genetic liability. These findings highlight the complex interplay between genetic risk of psychiatric disorders and behavioral traits in the context of SA, suggesting the need for a comprehensive mental health assessment for high-risk individuals.

## INTRODUCTION

Suicidal behaviors are a major health problem that results in approximately 700,000 deaths worldwide every year [1]. Suicide attempt (SA; self-initiated actions with the expectation of causing one’s own death) is a key predictor of death by suicide [2]. Notably, individuals affected by bipolar disorder (BD) and schizophrenia (SZ) have 17.1 and 12.9 higher odds of SA, respectively [3-5].

Identifying factors contributing to SA risk in individuals with BD and SZ is crucial for developing effective prevention and treatment strategies. Behavioral traits such as neuroticism, loneliness, and substance use have been associated with BD, SZ, and SA [6-8]. We hypothesize that these traits may affect SZ and BD association with SA. Mendelian randomization (MR) is an analytical approach that leverages genetic variants to estimate the causal effect of an exposure on an outcome of interest in observational data. While previous studies used a MR approach to estimate the causal effect of certain behavioral traits and socioeconomic factors (e.g., insomnia, smoking behavior, and educational attainment) on SA [9-12], many associations remain unexplored. These include behavioral traits related to SZ, BD, and many other important factors such as subjective well-being, trauma exposure, and risk-taking behavior.

To understand more comprehensively the network of associations linking BD and SZ to SA, we investigated their causal relationships with behavioral traits, socioeconomic factors, and substance use disorders utilizing large-scale genome-wide association data available from the Psychiatric Genomics Consortium (PGC) [13] and the UK Biobank (UKB) using a MR approach [14].

## MATERIALS AND METHODS

### Study overview

To evaluate the putative causal relationships among BD, SZ, behavioral traits, socioeconomic factors, substance use disorders, and SA, we used a genetically informed three-step design (Figure 1). First, we evaluated the effect of SZ and BD on SA. Then, we assessed the effect of BD and SZ on behavioral traits, socioeconomic factors, and substance use disorders. Finally, we evaluated the effect of these traits on SA. The associations were estimated using univariable and multivariable MR approaches.

**Figure 1.**
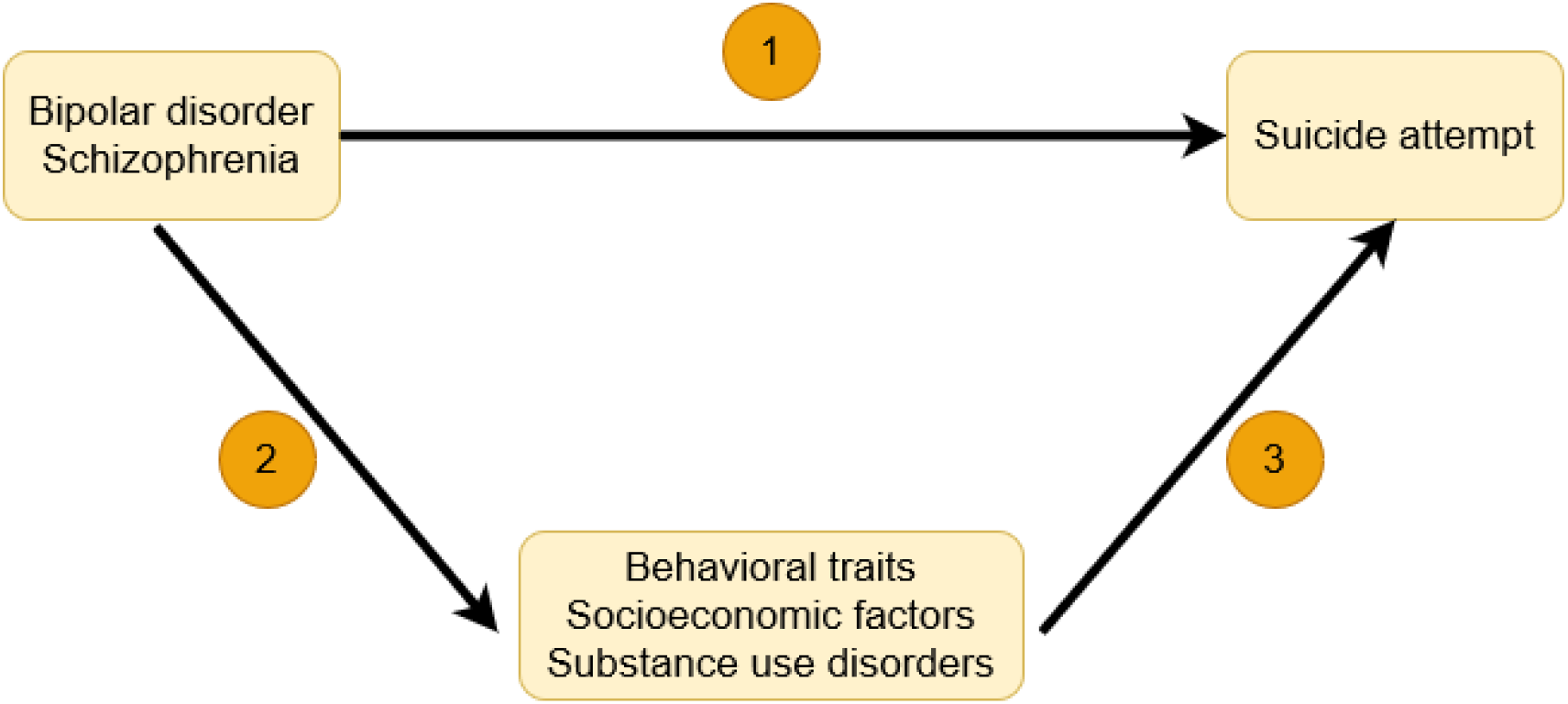
Study overview. Using univariable and multivariable MR approaches, we evaluated the total and direct effects of (1) bipolar disorder (BD) and schizophrenia (SZ) on suicide attempt (SA). Then, we evaluated (2) the putative causal effect of BD and SZ on behavioral traits, socioeconomic factors, and substance use disorders, and (3) the effect of these traits on SA.

### Data sources

Data from BD, SZ, and SA were derived from large-scale genome-wide association studies (GWAS) performed by the PGC [13]. The cohorts and GWAS procedures for BD (41,917 BD cases and 371,549 controls), SZ (53,386 cases and 77,258 controls), and SA (26,590 cases, 492,022 controls) have been previously described [15-17].

We evaluated the following behavioral traits: risk-taking behavior (n=466,571), insomnia (n=453,379), loneliness (n=445,024), cigarettes per day (n=618,489), drinks per week (n=2,428,851), neuroticism (n=170,911), subjective well-being (n=298,420), trauma exposure (n=24,094), mental distress (n=148,805), stress sensitivity (n=30,247), and alcohol use disorder identification test (AUDIT) total score (n=121,604) [18-26]. Additionally, because of the potential effect of socioeconomic factors and substance use disorders on BD-SZ-SA association, we also investigated household income (n=112,151), educational attainment (n=766,345), cannabis use disorder (n=357,806) and alcohol dependence (n=46,568) [27-30]. Details on data sources and a description of each trait are available in Supplementary Table 1. Briefly, genome-wide association statistics related to behavioral traits and socioeconomic factors were derived from the UKB cohort [14]. GWAS data related to cannabis use disorder and alcohol dependence were derived from PGC [13]. Because there is no large-scale GWAS for these phenotypes in other ancestry groups, the analyses were limited to datasets generated from individuals of European descent.

### Mendelian randomization

We estimated the putative causal effect (i.e., the sum of possible paths from the exposure on the outcome) among the phenotypes of interest using a two-sample MR approach [31]. This genetically informed causal inference framework is based on three assumptions: (i) the genetic instruments are associated with the outcome of interest; (ii) the genetic instruments are not associated with potential confounders of the risk factor–outcome association; and (iii) the genetic instruments affect the outcome only through their effect on the risk factor [32].

Because of the sample overlap among some of the datasets analyzed (i.e., GWAS including PGC and UKB samples), we calculated causal effects with the inverse variance weighted (IVW) method within the MRlap method, which is designed to correct the sample overlap among datasets used in two-sample MR analyses [33]. For each exposure, we defined a genetic instrument based on genome-wide significant variants (*P* < 5 × 10^−8^) that were linkage disequilibrium (LD)-independent (*r*^2^ < 0.001 within a 10 000-kilobase window) based on the 1000 Genomes Project Phase 3 reference panel for European populations [34]. When the number of LD-independent variants in the selected genetic instrument under these criteria was <10 single-nucleotide polymorphisms (SNPs), we also evaluated an alternative approach based on the inclusion of LD-independent variants available using more relaxed p-value thresholds. Specifically, we considered *P* < 5 × 10^−7^, *P* < 5 × 10^−6^, or 5 × 10^−5^ to include at least 10 LD-independent variants in each MR test (Supplementary Tables 3-11). As described below, we conducted multiple sensitivity analyses to identify possible violations of the MR assumptions and also applied multiple MR approaches that can account for possible assumption violations. These analyses were conducted using the R package TwoSampleMR [35].

We tested the presence of horizontal pleiotropy (variant affects the outcome indirectly, i.e., outside its direct effect on the exposure) and heterogeneity among the variants included in the genetic instruments using MR–Egger regression intercept [36] and heterogeneity tests [37], respectively. Furthermore, outliers contributing to the heterogeneity and horizontal pleiotropy within the genetic instruments were identified by leave-one-out analyses and the visual assessment of forest and funnel plots.

Unless otherwise noted, here we reported IVW effect estimates, as this method provides the highest statistical power [38]. As secondary analyses, we also used MR–Egger, weighted median, simple mode, and weighted mode approaches to verify the consistency in the direction and strength of the effect for each MR association with the IVW results [35]. To facilitate results interpretation and comparability across the evaluated traits, for all exposures, exposure effects on their respective outcomes were calculated as odds ratio (OR) for the outcome per one standard deviation (SD) increase in each exposure. For each MR test performed, we estimated *R*^2^ which corresponds to the proportion of variance of the exposure explained by the SNPs and mean F-statistic to evaluate the strength of the genetic instruments.

### Removing Sample Overlap among the Datasets

To further ensure the reliability of our results, we estimated the potential causal effect using data from non-overlapping samples. Because the sample size among non-overlapping datasets is much lower than the ones used in the full-scale analysis above, we evaluated the direction and strength of effect for each MR test rather than using p value thresholds.

When evaluating BD effect on SA, GWAS data from the PGC meta-analysis excluding the UKB for BD (20,352 BD cases and 31,358 controls) [39] were used as exposures, while the outcome dataset was the SA GWAS performed only in the UKB cohort (3,288 cases and 2,895 controls) [24]. Although the PGC SZ GWAS data did not include the UKB, we performed additional analyses on independent samples to replicate the direction and strength of the effects observed.

When testing the effect of behavioral traits and socioeconomic factors that were assessed in UKB, we used GWAS data from the PGC for BD (20,352 BD cases and 31,358 controls) [39], SA in individuals with BD (3,264 cases and 5,500 controls) [40], and SA in individuals with SZ (1,683 cases and 2,946 controls) [40]. Conversely, for cannabis use disorder and alcohol dependence that were evaluated in the PGC, we analyzed GWAS data for BD (1,257 BD cases and 369,930 controls) [41], SZ (701 SZ cases and 369,930 controls) [41], and SA (3,288 cases and 2,895 controls) [24] generated only from the UKB cohort. Details on data sources of non-overlapping samples are available in Supplementary Table 2.

### Multivariable Mendelian randomization

We estimated the direct causal effect of SZ, BD, behavioral traits, socioeconomic factors, and substance use disorders on SA and the independence of their effects using a multivariable MR (MVMR) approach [42]. Only traits that showed a significant putative effect on SA in the MR analysis with full samples (p-value < 0.05) were tested in a MVMR analysis. Since the MRlap approach does not permit to conduct multivariable investigations, we performed the MVMR analyses using the full-scale samples. Indeed, the estimates in the univariable MR analyses based on the full-scale samples were generally more conservative estimates than the ones generated using MRlap approach. This is due to the fact that sample overlap can bias MR estimates toward a null result. Accordingly, we considered significant effects as those reaching a nominal significance (p<0.05), because the MVMR analysis using the full-scale datasets is likely to generate conservative estimates. MVMR approach permitted us to estimate the independent effect of each exposure on the outcome of interest. The MVMR analysis was conducted using the MendelianRandomization R package [42].

## RESULTS

### Bipolar Disorder and Schizophrenia Effect on Suicide Attempt

In the two-sample MR analyses conducted using the MRlap approach, the genetic liabilities to BD and SZ were associated with SA (BD OR_MRlap_=1.24, p=3.88×10^−12^; SZ OR_MRlap_=1.09, p=2.44×10^−20^). These estimates were consistent with those obtained with the other MR methods considering full-scale and non-overlapping samples (Figure 2). Additionally, we did not observe heterogeneity or horizontal pleiotropy within genetic instruments (Supplementary Tables 3 and 4).

**Figure 2.**
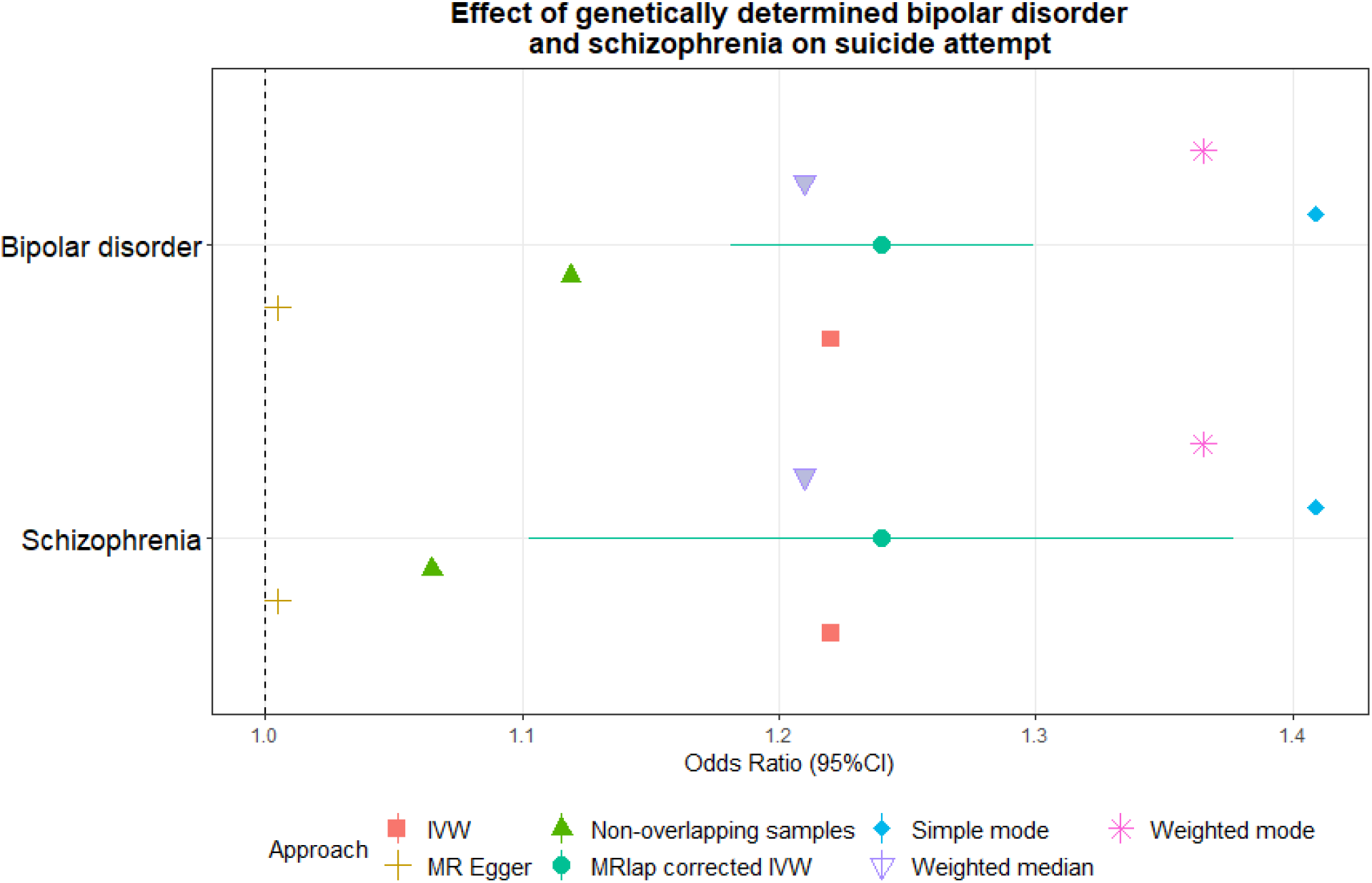
Effect of the genetic liability to schizophrenia and bipolar disorder on suicide attempt. Odds ratios were derived from MR estimates obtained from both full-scale and non-overlapping samples. 95% confidence intervals are reported for the MRlap estimates, considering potential sample overlap. Estimates from secondary MR methods are reported for each MR test. Abbreviations: inverse variance weighted (IVW).

### Schizophrenia and Bipolar Disorder Effect on Behavioral Traits, Socioeconomic Factors, and Substance Use Disorders

After Bonferroni multiple testing correction (p<0.003), we found that the genetic liability to BD was associated with educational attainment (OR_MRlap_=1.31, p=6.55×10^−6^), risk-taking behavior (p=0.02; OR_MRlap_=1.24, p=1.88×10^−6^), and mental distress (OR_MRlap_=1.42, p=7.15×10^−10^).

Considering a Bonferroni correction for the number of traits tested (p<0.003), the genetic liability to SZ was associated with risk-taking behavior (OR_IVW_=1.02, p=1.99×10^−8^), trauma exposure (OR_IVW_=1.10, p=0.002), mental distress (OR_IVW_=1.06, p=3.57×10^−7^), household income (OR_IVW_=0.99, p=0.001), subjective well-being (OR_MRlap_=0.94, p=2.7×10^−7^), loneliness (OR_IVW_=1.004, p=0.003), neuroticism (OR_MRlap_=1.09, p=3.65×10^−5^), drinks per week (OR_IVW_=1.01, p=4.12×10^−4^), alcohol dependence (OR_MRlap_=1.14, p=1.07×10^−5^), and cannabis use disorder (OR_MRlap_=1.08, p=1.36×10^−12^). For the MR tests that did not have any sample overlap between exposure and outcome datasets, we considered IVW estimates to describe SZ effects.

In most of the cases, Bonferroni-significant MRlap estimates were consistent with the estimates obtained from the other MR methods considering full-scale and non-overlapping samples (Figure 3). Additionally, we did not observe evidence of heterogeneity or horizontal pleiotropy among genetic instruments related to BD (Supplemental Tables 5 and 6) and SZ (Supplemental Tables 7 and 8).

**Figure 3.**
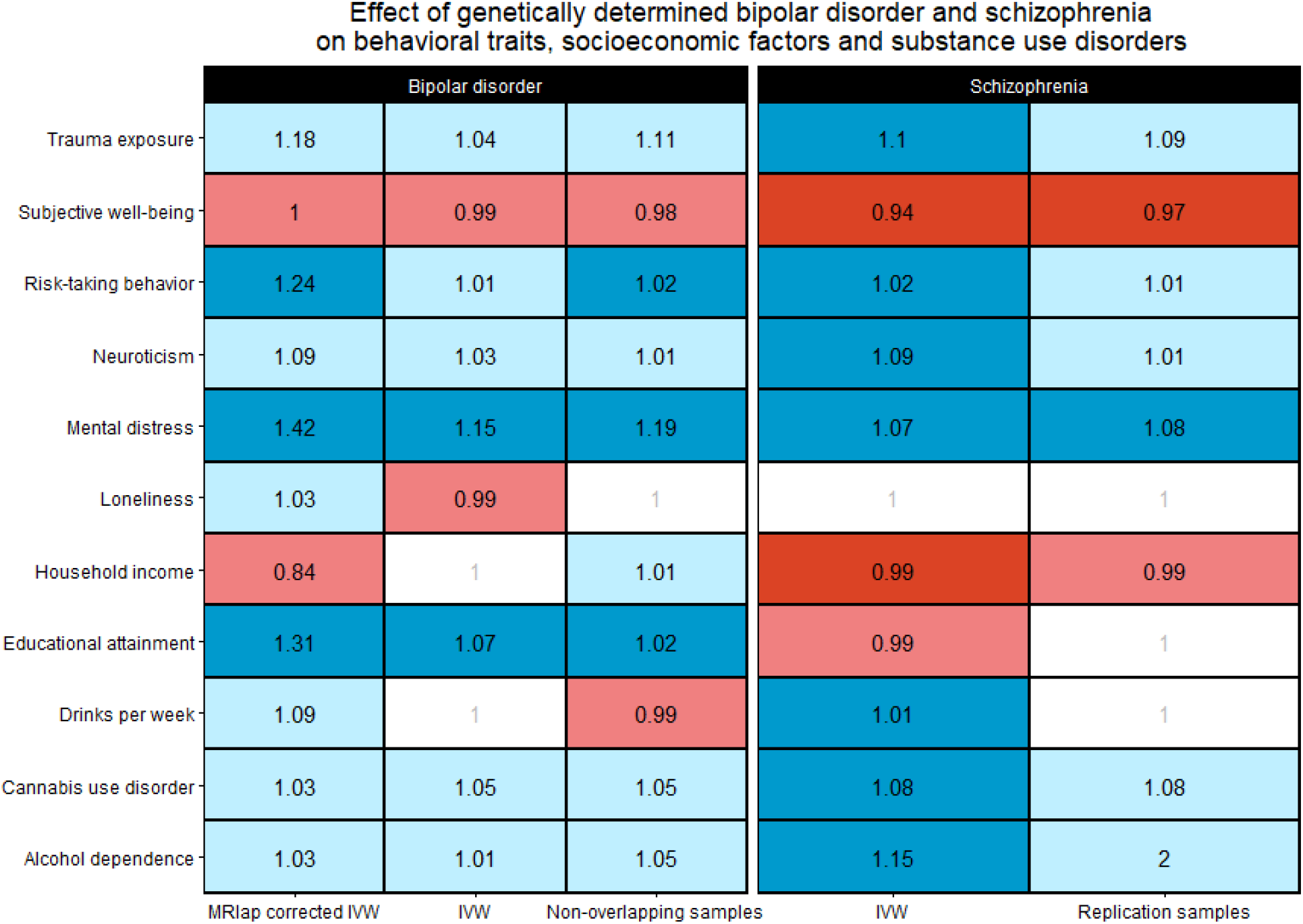
Consistency of odds ratios across Mendelian randomization methods and data sources (i.e., full-scale and non-overlapping samples) considering associations of bipolar disorder and schizophrenia genetic liabilities with behavioral traits, socioeconomic factors and substance use disorders. Only traits with Bonferroni significant estimates in the full-scale samples in either bipolar disorder (MRlap corrected IVW) or schizophrenia (IVW) are shown in the Figure. Bonferroni significant estimates (p<0.003) are highlighted in blue for positive effects and red for negative effects. Light blue and red indicate positive and negative effects that did not survive multiple testing correction, respectively. Cells reporting non-significant association with odds ratio <1.01 were indicated in white. Abbreviations: inverse variance weighted (IVW).

### Effect of Behavioral and Psychiatric Traits, Socioeconomic Factors, and Substance Use Disorders on Suicide Attempt

With respect to behavioral traits, MRlap approach showed Bonferroni-significant effects of risk-taking behavior (OR_MRlap_=1.32, p=6.19×10^−5^), mental distress (OR_MRlap_=1.30, p=0.001), and loneliness (OR_MRlap_=1.33, p=0.002) on SA. Conversely, educational attainment showed a protective association with SA (OR_MRlap_=0.89, p=6.34×10^−11^). Risk-taking behavior, loneliness, and educational attainment associations with SA were consistent across MR methods in the full-scale data and in at least one of the non-overlapping SA samples (Figure 4). Additionally, we did not observe evidence of horizontal pleiotropy and heterogeneity among the genetic instruments related to the phenotypes (Supplementary Tables 9, 10, and 11).

**Figure 4.**
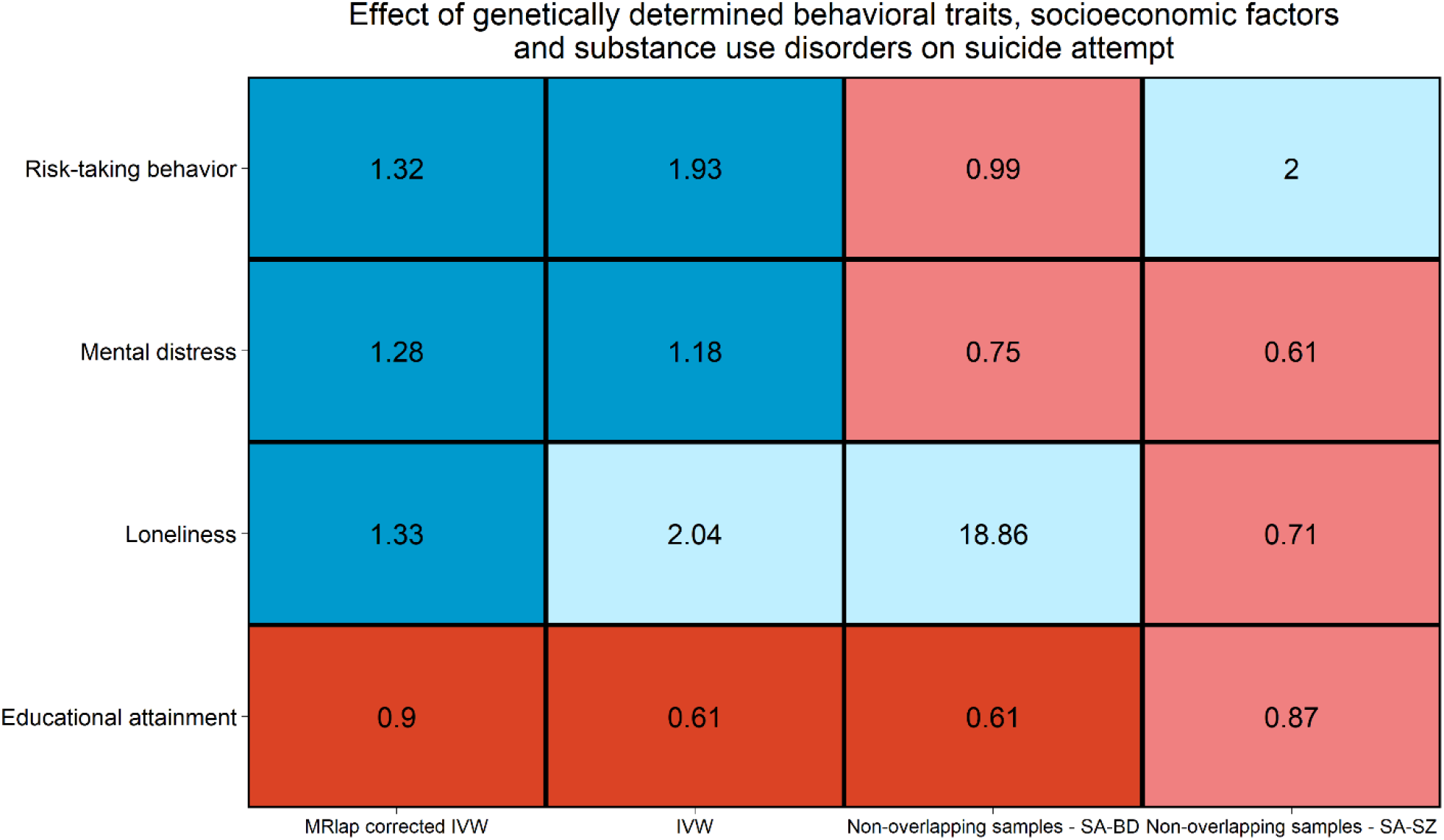
Effect consistency across Mendelian randomization methods and data sources (i.e., full-scale and non-overlapping samples) considering associations of genetic liability for behavioral traits, socioeconomic factors and substance use disorders with suicide-attempt identified as Bonferroni significant in the MRlap analysis. Bonferroni significant estimates (p<0.003) are indicated in blue for positive effects and red for negative effects. Light blue and red indicate positive and negative effects that did not survive multiple testing correction, respectively. Abbreviations: inverse variance weighted (IVW); Suicide attempt in individuals with bipolar disorder (SA-BD); suicide attempt in individuals with schizophrenia (SA-SZ).

### Multivariable Mendelian randomization

The MVMR analysis showed that BD and SZ genetic liabilities have independent effects on SA (BD: OR_MVMR_=1.24, p=6.5×10^−10^; SZ: OR_MVMR_=1.07, p=0.002; Figure 5). Additionally, the positive effect of mental distress (OR_MVMR_=1.17, p=1.02×10^−4^) and risk-taking behavior (OR_MVMR_=1.52, p=0.028) on SA remained significant after accounting for SZ. Conversely, the effect of both these psychiatric traits on SA was null when accounting for the effect of BD (Supplementary Table 12). Also, the protective effect of educational attainment on SA (OR_MVMR_=0.63, p=2.97×10^−15^) was independent of BD genetic liability, while the effect of loneliness on SA was null after accounting for SZ genetic liability (Supplementary Table 12). We observed significant heterogeneity (p=1.43×10^−7^) among the genetic instruments related to BD and mental distress that remained significant after removal of potential outliers. We did not observe evidence of horizontal pleiotropy and heterogeneity among the genetic instruments related to the rest of phenotypes (Supplementary Table 12).

**Figure 5.**
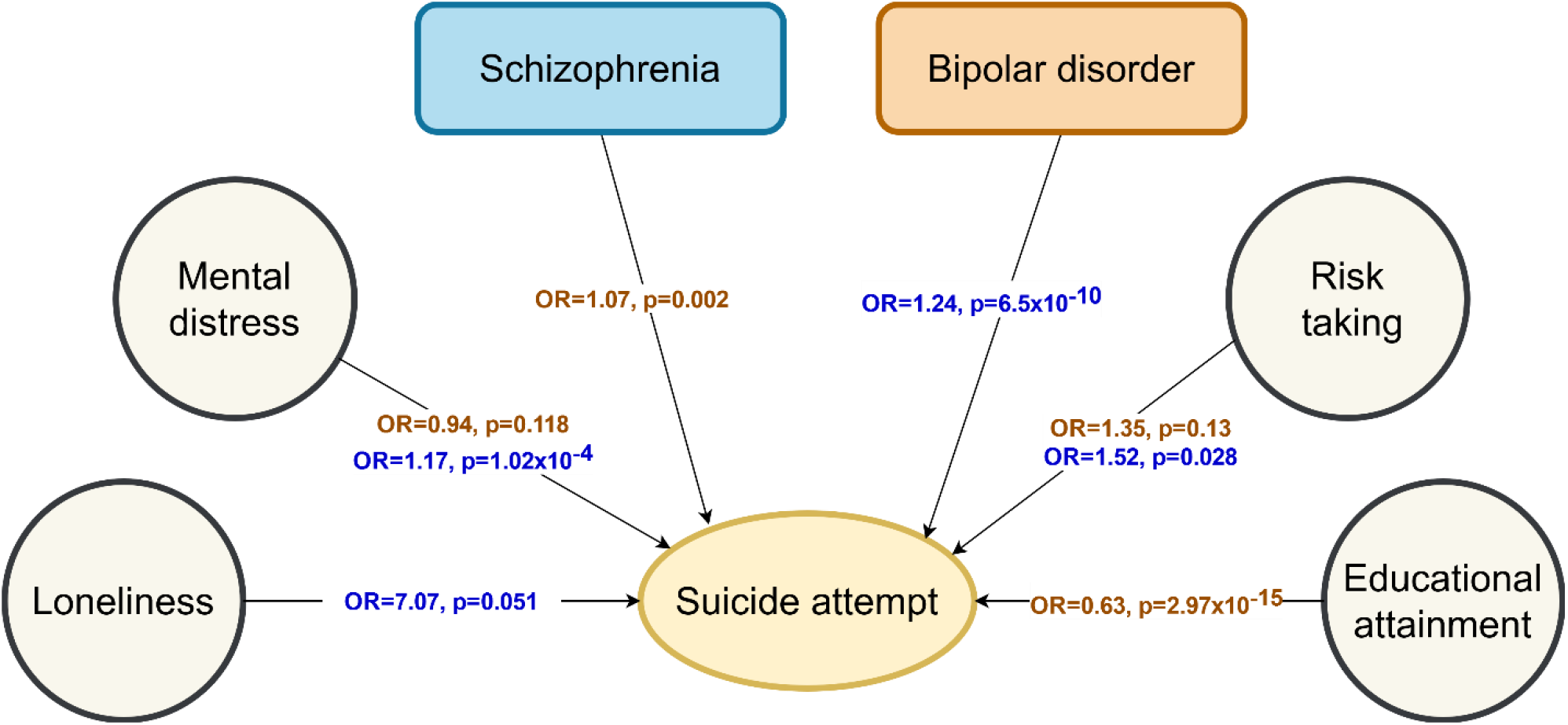
Multivariable Mendelian Randomization results. Arrows indicate the direction of the effect between the traits. ORs and p-values from each association are shown in the middle of the arrows for the multivariable MR analysis accounting for bipolar disorder (in brown) and schizophrenia (in blue). Only traits that showed a significant putative effect on suicide attempt in the two-sample Mendelian Randomization analysis with full samples (p-value < 0.05) were tested in the multivariable Mendelian Randomization analysis. Abbreviations: Odds ratio (OR); p-value (p).

## DISCUSSION

While previous analyses showed that genetic liabilities to BD and SZ increase the odds of suicidal behaviors [43, 44], the present study expanded the understanding of the dynamics underlying these associations, demonstrating that their effects on SA are independent of each other and of several behavioral traits, socioeconomic factors, and substance use disorders.

A noteworthy finding was the effect of BD and SZ genetic liabilities on risk-taking behavior and mental distress, which then are associated with SA. There is substantial genetic overlap among BD, SZ, and risk-taking behavior [45]. Observational studies have confirmed the role of risk-taking behavior and mental distress in increasing the risk of suicide-related outcomes [46,47]. A theoretical suicide model has highlighted that risk-taking behavior serves as a potential mediator between mental distress and SAs, implying that engaging in risky activities may contribute to the transition from mental distress to suicidal behavior [48]. With respect to mental disorders, our MVMR analyses suggest that the effect of mental distress and risk-taking behavior on SA is independent of SZ genetic liability, but not of BD. This finding highlights the disorder-specific relevance of both behavioral traits for BD, particularly risk-taking behavior, for which higher levels of risk-taking behavior in patients with BD with respect to patients with SZ have been reported [49]. Also, this finding raises the possibility that shared risk factors for SA might have disorder-specific mechanisms that contribute to increased suicide risk. Furthermore, our results highlight the role of risk-taking behavior and mental distress as potential mediators in the association between BD/SZ and SA. Such findings encourage future research to account for these traits, as they might act as potential confounders when investigating the genetic association between BD/SZ diagnoses and SA. The identification of mental distress and risk-taking behavior as contributors to the risk of SA underscores their potential as promising targets for assessment and interventional programs in patients with mental disorders. Accordingly, during the treatment of individuals with BD, a focused approach toward assessing and addressing these behavioral traits may be emphasized. Furthermore, for individuals with SZ, these assessments and interventions should be continued even during periods of resolution and remission of SZ symptoms, to effectively reduce SA risk.

The effect of SZ on loneliness is consistent with previous reports [50,51], but loneliness association with SA has not been previously reported by previous genetically informed causal inference analyses. The interpersonal theory of suicide indicates that mental disorders can increase the risk of experiencing social isolation and loneliness, contributing to hopelessness [52]. These factors can further impede individuals from seeking and receiving support, thereby increasing their vulnerability to suicide [53,54]. Our MVMR analysis highlighted that the effect of loneliness on SA is not independent of SZ. The effect of loneliness on SA in the context of SZ might be influenced by the association of loneliness with multiple comorbidities such as substance use, major depression, and anxiety in individuals with high SZ genetic risk [55, 56]. This finding also highlights the need for interventions targeting loneliness in individuals with SZ and people with SZ family history. By addressing loneliness, we may potentially reduce SA in this high-risk group. These findings increase the understanding of the link among SZ, loneliness, and SA, providing novel information that can contribute to the development of targeted interventions aimed at fostering social connectedness to reduce SA risk in vulnerable individuals. BD’s effect on educational attainment and the association of both with SA are consistent with previous MR studies [10, 15, 57]. Additionally, the direct effect of BD and educational attainment on SA identified by the present MVMR analysis is in line with a previous study [10]. Also, the lack of association of SZ with educational attainment is in line with a previous report [57]. This may be due to the complex relationship between SZ and educational attainment where many individual loci across the genome appear to have discordant pleiotropic effects with these phenotypes [58].

In addition to these associations, we identified possible cause-effect relationships linking SZ genetic liabilities to behavioral traits, socioeconomic factors, and substance use disorders that appeared to directly affect SA. Although they may not be directly implicated in suicidality, they can help to understand the dynamics affecting SZ patients and individuals experiencing psychotic symptoms. For example, the association of SZ genetic liability with reduced odds of subjective well-being and household income highlights the importance of mental health support as a preventive strategy for enhancing the well-being and the economic security of individuals with SZ and also of individuals with high SZ genetic risk such as family members of SZ patients [59, 60]. Similarly, we observed that SZ genetic liability increased the odds of neuroticism. This is a personality trait associated with many negative outcomes related to both mental and physical health [61]. While to our knowledge, no previous MR analysis explored this relationship, there are multiple studies reporting high neuroticism scores among SZ patients [62, 63]. Our result indicates a specific direction in this relationship that could support the development of personality screening programs among SZ patients to reduce the negative consequences of this mental illness.

In our study, we confirmed the effect of SZ on alcohol dependence and cannabis use disorder. These associations are supported by multiple epidemiological and genetically informed studies [15, 64-67]. However, with respect to alcohol-related phenotypes, we did not observe any SZ association with AUDIT total score. There is growing literature highlighting the distinct genetic profiles between alcohol consumption and alcohol use disorder [68, 69]. Additionally, previous MR studies showed that the genetic liability of other psychiatric disorders (e.g., major depression) could lead to increased odds of alcohol dependence but not of alcohol consumption [70]. In line with these previous findings, the current results support that SZ genetic liability may have a causal effect on alcohol-related pathologic behaviors, but not on traits more closely related to general alcohol consumption.

Several limitations should be acknowledged in the present study. First, the analyses were conducted on datasets generated from individuals of European descent. This may limit the generalizability of our results to populations with different ancestral origins. Future research should include diverse ancestral backgrounds to evaluate the effect of BD, SZ, and related phenotypes on SA more comprehensively. Second, despite conducting multiple sensitivity analyses, the influence of potential confounding variables on our results cannot be entirely ruled out. Complementary studies with robust control measures are needed to confirm and further explore the reported findings. Third, although we used multiple approaches to account for the sample overlap, we cannot completely exclude possible biases. Additionally, some of the null results observed may be due to the limited statistical power of the genome-wide information analyzed. Future studies investigating causal associations between other mental disorders, such as major depression disorder and generalized anxiety disorder with SA are essential to improve our understanding of the mechanisms involved in the association between mental disorders and SA. Lastly, it is important to note that the evaluated traits are multifactorial in nature and influenced by factors other than genetics, which was the primary focus of this study.

In conclusion, our study highlights the complex network of associations linking BD, SZ, and several related phenotypes to SA. Specifically, novel putative associations were identified for subjective well-being, risk-taking behavior, loneliness, and mental distress, which are relevant by being influenced by BD/SZ and also having a putative effect on SA. Furthermore, it is crucial to pay special attention to disorder-specific causal associations, such as the putative effect of SZ on loneliness, as they are particularly relevant for the development of targeted preventive strategies. Thus, therapeutical and assessment measures targeting the complex interplay among mental illnesses, behavioral traits, and socioeconomic factors could help reduce suicide risk among vulnerable individuals such as those with high BD and SZ genetic risk.

## Data Availability

All data produced in the present work are contained in the manuscript

## ACKNOWLEDGEMENTS

The authors thank the participants and the investigators involved in the Psychiatric Genomics Consortium and the UK Biobank.

## AUTHOR CONTRIBUTIONS

BCM and RP conceived and designed the study. BCM and NA performed the analysis. GRF, ARD, CWB contributed to the interpretation of the results. BCM and RP drafted the manuscript. All authors edited or approved the final manuscript and are in agreement to be accountable for its contents.

## REFERENCES

1. World Health Organization. Live life: an implementation guide for suicide prevention in countries. World Health Organization; 2021.

2. American Psychiatric Association. Diagnostic and Statistical Manual of Mental Disorders. Fifth Edition. American Psychiatric Association; 2013.

3. Dome P, Rihmer Z, Gonda X. Suicide Risk in Bipolar Disorder: A Brief Review. Medicina (Kaunas). 2019;55:403.

4. Pompili M, Amador XF, Girardi P, Harkavy-Friedman J, Harrow M, Kaplan K, et al. Suicide risk in schizophrenia: learning from the past to change the future. Ann Gen Psychiatry. 2007;6:10.

5. Chesney E, Goodwin GM, Fazel S. Risks of all-cause and suicide mortality in mental disorders: a meta-review. World Psychiatry. 2014;13:153–160.

6. Shaw RJ, Cullen B, Graham N, Lyall DM, Mackay D, Okolie C, et al. Living alone, loneliness and lack of emotional support as predictors of suicide and self-harm: A nine-year follow up of the UK Biobank cohort. Journal of Affective Disorders. 2021;279:316–323.

7. Hafferty JD, Navrady LB, Adams MJ, Howard DM, Campbell AI, Whalley HC, et al. The role of neuroticism in self-harm and suicidal ideation: results from two UK population-based cohorts. Soc Psychiatry Psychiatr Epidemiol. 2019;54:1505–1518.

8. Poorolajal J, Haghtalab T, Farhadi M, Darvishi N. Substance use disorder and risk of suicidal ideation, suicide attempt and suicide death: a meta-analysis. J Public Health. 2016;38:e282–e291.

9. Nassan M, Daghlas I, Winkelman JW, Dashti HS, International Suicide Genetics Consortium, Saxena R. Genetic evidence for a potential causal relationship between insomnia symptoms and suicidal behavior: a Mendelian randomization study. Neuropsychopharmacol. 2022;47:1672–1679.

10. Rosoff DB, Kaminsky ZA, McIntosh AM, Davey Smith G, Lohoff FW. Educational attainment reduces the risk of suicide attempt among individuals with and without psychiatric disorders independent of cognition: a bidirectional and multivariable Mendelian randomization study with more than 815,000 participants. Transl Psychiatry. 2020;10:388.

11. Harrison R, Munafò MR, Davey Smith G, Wootton RE. Examining the effect of smoking on suicidal ideation and attempts: triangulation of epidemiological approaches. Br J Psychiatry. 2020;217:701–707.

12. Kootbodien T, London L, Martin LJ, Defo J, Ramesar R. The shared genetic architecture of suicidal behaviour and psychiatric disorders: A genomic structural equation modelling study. Front Genet. 2023;14:1083969.

13. Sullivan PF, Agrawal A, Bulik CM, Andreassen OA, Børglum AD, Breen G, et al. Psychiatric Genomics: An Update and an Agenda. Am J Psychiatry. 2018;175:15–27.

14. Bycroft C, Freeman C, Petkova D, Band G, Elliott LT, Sharp K, et al. The UK Biobank resource with deep phenotyping and genomic data. Nature. 2018;562:203–209.

15. Mullins N, Forstner AJ, O’Connell KS, Coombes B, Coleman JRI, Qiao Z, et al. Genome-wide association study of more than 40,000 bipolar disorder cases provides new insights into the underlying biology. Nat Genet. 2021;53:817–829.

16. Trubetskoy V, Pardiñas AF, Qi T, Panagiotaropoulou G, Awasthi S, Bigdeli TB, et al. Mapping genomic loci implicates genes and synaptic biology in schizophrenia. Nature. 2022;604:502–508.

17. Mullins N, Kang J, Campos AI, Coleman JRI, Edwards AC, Galfalvy H, et al. Dissecting the Shared Genetic Architecture of Suicide Attempt, Psychiatric Disorders, and Known Risk Factors. Biol Psychiatry. 2022;91:313–327.

18. Karlsson Linnér R, Biroli P, Kong E, Meddens SFW, Wedow R, Fontana MA, et al. Genome-wide association analyses of risk tolerance and risky behaviors in over 1 million individuals identify hundreds of loci and shared genetic influences. Nat Genet. 2019;51:245–257.

19. Lane JM, Jones SE, Dashti HS, Wood AR, Aragam KG, van Hees VT, et al. Biological and clinical insights from genetics of insomnia symptoms. Nat Genet. 2019;51:387–393.

20. Day FR, Ong KK, Perry JRB. Elucidating the genetic basis of social interaction and isolation. Nat Commun. 2018;9:2457.

21. Saunders GRB, Wang X, Chen F, Jang S-K, Liu M, Wang C, et al. Genetic diversity fuels gene discovery for tobacco and alcohol use. Nature. 2022;612:720–724.

22. Okbay A, Baselmans BML, De Neve J-E, Turley P, Nivard MG, Fontana MA, et al. Genetic variants associated with subjective well-being, depressive symptoms, and neuroticism identified through genome-wide analyses. Nat Genet. 2016;48:624–633.

23. Coleman JRI, Peyrot WJ, Purves KL, Davis KAS, Rayner C, Choi SW, et al. Genome-wide gene-environment analyses of major depressive disorder and reported lifetime traumatic experiences in UK Biobank. Mol Psychiatry. 2020;25:1430–1446.

24. Jiang L, Zheng Z, Fang H, Yang J. A generalized linear mixed model association tool for biobank-scale data. Nat Genet. 2021;53:1616–1621.

25. Arnau-Soler A, Adams MJ, Generation Scotland, Major Depressive Disorder Working Group of the Psychiatric Genomics Consortium, Hayward C, Thomson PA. Genome-wide interaction study of a proxy for stress-sensitivity and its prediction of major depressive disorder. PLoS One. 2018;13:e0209160.

26. Sanchez-Roige S, Palmer AA, Fontanillas P, Elson SL, 23andMe Research Team, the Substance Use Disorder Working Group of the Psychiatric Genomics Consortium, Adams MJ, et al. Genome-Wide Association Study Meta-Analysis of the Alcohol Use Disorders Identification Test (AUDIT) in Two Population-Based Cohorts. Am J Psychiatry. 2019;176:107–118.

27. Hill WD, Davies NM, Ritchie SJ, Skene NG, Bryois J, Bell S, et al. Genome-wide analysis identifies molecular systems and 149 genetic loci associated with income. Nat Commun. 2019;10:5741.

28. 23andMe Research Team, COGENT (Cognitive Genomics Consortium), Social Science Genetic Association Consortium, Lee JJ, Wedow R, Okbay A, et al. Gene discovery and polygenic prediction from a genome-wide association study of educational attainment in 1.1 million individuals. Nat Genet. 2018;50:1112–1121.

29. Johnson EC, Demontis D, Thorgeirsson TE, Walters RK, Polimanti R, Hatoum AS, et al. A large-scale genome-wide association study meta-analysis of cannabis use disorder. Lancet Psychiatry. 2020;7:1032–1045.

30. Walters RK, Polimanti R, Johnson EC, McClintick JN, Adams MJ, Adkins AE, et al. Transancestral GWAS of alcohol dependence reveals common genetic underpinnings with psychiatric disorders. Nat Neurosci. 2018;21:1656–1669.

31. Sanderson E, Glymour MM, Holmes MV, Kang H, Morrison J, Munafò MR, et al. Mendelian randomization. Nat Rev Methods Primers. 2022;2:6.

32. de Leeuw C, Savage J, Bucur IG, Heskes T, Posthuma D. Understanding the assumptions underlying Mendelian randomization. Eur J Hum Genet. 2022;30:653–660.

33. Mounier N, Kutalik Z. Bias correction for inverse variance weighting Mendelian randomization. Genet Epidemiol. 2023;47:314–331.

34. 1000 Genomes Project Consortium, Auton A, Brooks LD, Durbin RM, Garrison EP, Kang HM, et al. A global reference for human genetic variation. Nature. 2015;526:68–74.

35. Hemani G, Zheng J, Elsworth B, Wade KH, Haberland V, Baird D, et al. The MR-Base platform supports systematic causal inference across the human phenome. eLife. 2018;7:e34408.

36. Bowden J, Davey Smith G, Burgess S. Mendelian randomization with invalid instruments: effect estimation and bias detection through Egger regression. International Journal of Epidemiology. 2015;44:512–525.

37. Burgess S, Bowden J, Fall T, Ingelsson E, Thompson SG. Sensitivity Analyses for Robust Causal Inference from Mendelian Randomization Analyses with Multiple Genetic Variants. Epidemiology. 2017;28:30–42.

38. Hemani G, Bowden J, Davey Smith G. Evaluating the potential role of pleiotropy in Mendelian randomization studies. Human Molecular Genetics. 2018;27:R195–R208.

39. eQTLGen Consortium, BIOS Consortium, the Bipolar Disorder Working Group of the Psychiatric Genomics Consortium, Stahl EA, Breen G, Forstner AJ, et al. Genome-wide association study identifies 30 loci associated with bipolar disorder. Nat Genet. 2019;51:793–803.

40. Mullins N, Bigdeli TB, Børglum AD, Coleman JRI, Demontis D, Mehta D, et al. GWAS of Suicide Attempt in Psychiatric Disorders and Association With Major Depression Polygenic Risk Scores. AJP. 2019;176:651–660.

41. Pan UKBB | Pan UKBB. https://pan.ukbb.broadinstitute.org/. Accessed 17 March 2023.

42. Burgess S, Thompson SG. Multivariable Mendelian randomization: the use of pleiotropic genetic variants to estimate causal effects. Am J Epidemiol. 2015;181:251–260.

43. Mullins N, Forstner AJ, O’Connell KS, Coombes B, Coleman JRI, Qiao Z, et al. Genome-wide association study of more than 40,000 bipolar disorder cases provides new insights into the underlying biology. Nat Genet. 2021;53:817–829.

44. Lim KX, Rijsdijk F, Hagenaars SP, Socrates A, Choi SW, Coleman JRI, et al. Studying individual risk factors for self-harm in the UK Biobank: A polygenic scoring and Mendelian randomisation study. PLoS Med. 2020;17:e1003137.

45. Hindley G, Bahrami S, Steen NE, O’Connell KS, Frei O, Shadrin A, et al. Characterising the shared genetic determinants of bipolar disorder, schizophrenia and risk-taking. Transl Psychiatry. 2021;11:466.

46. Athey A, Overholser J, Bagge C, Dieter L, Vallender E, Stockmeier CA. Risk-taking behaviors and stressors differentially predict suicidal preparation, non-fatal suicide attempts, and suicide deaths. Psychiatry Res. 2018;270:160–167.

47. Bell S, Russ TC, Kivimäki M, Stamatakis E, Batty GD. Dose-Response Association Between Psychological Distress and Risk of Completed Suicide in the General Population. JAMA Psychiatry. 2015;72:1254.

48. O’Connor RC, Kirtley OJ. The integrated motivational-volitional model of suicidal behaviour. Philos Trans R Soc Lond B Biol Sci. 2018;373:20170268.

49. Reddy LF, Lee J, Davis MC, Altshuler L, Glahn DC, Miklowitz DJ, et al. Impulsivity and Risk Taking in Bipolar Disorder and Schizophrenia. Neuropsychopharmacol. 2014;39:456–463.

50. Andreu-Bernabeu Á, Díaz-Caneja CM, Costas J, De Hoyos L, Stella C, Gurriarán X, et al. Polygenic contribution to the relationship of loneliness and social isolation with schizophrenia. Nat Commun. 2022;13:51.

51. Zhang F, Baranova A, Zhou C, Cao H, Chen J, Zhang X, et al. Causal influences of neuroticism on mental health and cardiovascular disease. Hum Genet. 2021;140:1267–1281.

52. Chu C, Buchman-Schmitt JM, Stanley IH, Hom MA, Tucker RP, Hagan CR, et al. The interpersonal theory of suicide: A systematic review and meta-analysis of a decade of cross-national research. Psychol Bull. 2017;143:1313–1345.

53. Van Orden KA, Witte TK, Cukrowicz KC, Braithwaite SR, Selby EA, Joiner TE. The interpersonal theory of suicide. Psychological Review. 2010;117:575–600.

54. Lim KX, Oginni OA, Rimfeld K, Pingault J-B, Rijsdijk F. Investigating the causal risk factors for self-harm by integrating Mendelian randomisation within twin modelling. Behav Genet. 2022;52:324–337.

55. Trémeau F, Antonius D, Malaspina D, Goff DC, Javitt DC. Loneliness in schizophrenia and its possible correlates. An exploratory study. Psychiatry Research. 2016;246:211–217.

56. Eglit GML, Palmer BW, Martin AS, Tu X, Jeste DV. Loneliness in schizophrenia: Construct clarification, measurement, and clinical relevance. PLoS ONE. 2018;13:e0194021.

57. Demange PA, Boomsma DI, Van Bergen E, Nivard MG. Evaluating the causal relationship between educational attainment and mental health. Psychiatry and Clinical Psychology; 2023.

58. Lam M, Hill WD, Trampush JW, Yu J, Knowles E, Davies G, et al. Pleiotropic Meta-Analysis of Cognition, Education, and Schizophrenia Differentiates Roles of Early Neurodevelopmental and Adult Synaptic Pathways. Am J Hum Genet. 2019;105:334–350.

59. He Y, Tanaka A, Kishi T, Li Y, Matsunaga M, Tanihara S, et al. Recent findings on subjective well-being and physical, psychiatric, and social comorbidities in individuals with schizophrenia: A literature review. Neuropsychopharmacol Rep. 2022;42:430–436.

60. Chong HY, Teoh SL, Wu DB-C, Kotirum S, Chiou C-F, Chaiyakunapruk N. Global economic burden of schizophrenia: a systematic review. Neuropsychiatr Dis Treat. 2016;12:357–373.

61. Widiger TA, Oltmanns JR. Neuroticism is a fundamental domain of personality with enormous public health implications. World Psychiatry. 2017;16:144–145.

62. Franquillo AC, Guccione C, Angelini G, Carpentieri R, Ducci G, Caretti V. The Role of Personality in Schizophrenia and Psychosis: A Systematic Review. Clin Neuropsychiatry. 2021;18:28–40.

63. Shi J, Yao Y, Zhan C, Mao Z, Yin F, Zhao X. The Relationship Between Big Five Personality Traits and Psychotic Experience in a Large Non-clinical Youth Sample: The Mediating Role of Emotion Regulation. Front Psychiatry. 2018;9:648.

64. Vaucher J, Keating BJ, Lasserre AM, Gan W, Lyall DM, Ward J, et al. Cannabis use and risk of schizophrenia: a Mendelian randomization study. Mol Psychiatry. 2018;23:1287–1292.

65. Johnson EC, Hatoum AS, Deak JD, Polimanti R, Murray RM, Edenberg HJ, et al. The relationship between cannabis and schizophrenia: a genetically informed perspective. Addiction. 2021;116:3227–3234.

66. Orri M, Séguin JR, Castellanos-Ryan N, Tremblay RE, Côté SM, Turecki G, et al. A genetically informed study on the association of cannabis, alcohol, and tobacco smoking with suicide attempt. Mol Psychiatry. 2021;26:5061–5070.

67. Polimanti R, Agrawal A, Gelernter J. Schizophrenia and substance use comorbidity: a genome-wide perspective. Genome Med. 2017;9:25.

68. Kranzler HR, Zhou H, Kember RL, Vickers Smith R, Justice AC, Damrauer S, et al. Genome-wide association study of alcohol consumption and use disorder in 274,424 individuals from multiple populations. Nat Commun. 2019;10:1499.

69. Gelernter J, Polimanti R. Genetics of substance use disorders in the era of big data. Nat Rev Genet. 2021;22:712–729.

70. Polimanti R, Peterson RE, Ong J-S, MacGregor S, Edwards AC, Clarke T-K, et al. Evidence of causal effect of major depression on alcohol dependence: findings from the psychiatric genomics consortium. Psychol Med. 2019;49:1218–1226.

